# A throughput serological Western blot system using whole virus lysate for the concomitant detection of antibodies against SARS-CoV-2 and human endemic Coronaviridae

**DOI:** 10.1101/2020.07.31.20165019

**Authors:** Simon Fink, Felix Ruoff, Aaron Stahl, Matthias Becker, Philipp Kaiser, Bjoern Traenkle, Daniel Junker, Frank Weise, Natalia Ruetalo, Sebastian Hörber, Andreas Peter, Annika Nelde, Juliane Walz, Gérard Krause, Katja Schenke-Layland, Thomas Joos, Ulrich Rothbauer, Nicole Schneiderhan-Marra, Michael Schindler, Markus F. Templin

**Author notes:** Corresponding author: Dr. Markus Templin. These authors contributed equally to this work (Simon Fink, Felix Ruoff).

## Abstract

**Background:** Seroreactivity against human endemic coronaviruses has been linked to disease severity after SARS-CoV-2 infection. Assays that are capable of concomitantly detecting antibodies against endemic coronaviridae such as OC43, 229E, NL63, and SARS-CoV-2 may help to elucidate this question. We set up a platform for serum-screening and developed a bead-based Western blot system, namely DigiWest, capable of running hundreds of assays using microgram amounts of protein prepared directly from different viruses.

**Methods:** The parallelized and miniaturised DigiWest assay was adapted for detecting antibodies using whole protein extract prepared from isolated SARS-CoV-2 virus particles. After characterisation and optimization of the newly established test, whole virus lysates of OC43, 229E, and NL63 were integrated into the system.

**Results:** The DigiWest-based immunoassay system for detection of SARS-CoV-2 specific antibodies shows a sensitivity of 87.2 % and diagnostic specificity of 100 %. Concordance analysis with the SARS-CoV-2 immunoassays available by Roche, Siemens, and Euroimmun indicates a comparable assay performance (Cohen’s Kappa ranging from 0.8799-0.9429). In the multiplexed assay, antibodies against the endemic coronaviruses OC43, 229E, and NL63 were detected, displaying a high incidence of seroreactivity against these coronaviruses.

**Conclusion:** The DigiWest-based immunoassay, which uses authentic antigens from isolated virus particles, is capable of detecting individual serum responses against SARS-CoV-2 with high specificity and sensitivity in one multiplexed assay. It shows high concordance with other commercially available serologic assays. The DigiWest approach enables a concomitant detection of antibodies against different endemic coronaviruses and will help to elucidate the role of these possibly cross-reactive antibodies.

## Introduction

Severe acute respiratory syndrome coronavirus 2 (SARS-CoV-2) is a newly identified beta coronavirus that crossed the species barrier and found its way into the human population in 2019. It causes the coronavirus disease 2019 (COVID-19), and the ongoing pandemic has a devastating effect on wide parts of the human population (1). The virus is highly contagious causing the disease to spread very rapidly, yet symptoms of infected individuals vary widely. A fraction of COVID-19 patients develops a fatal course of the disease, while mild COVID-19 cases are frequently observed (2). Different co-morbidity factors were recently identified, whereas the prediction of the course of the disease is not yet possible (3). Protective antibodies formed after infection are associated with viral clearance, but the occurrence of high antibody titres has also been linked to more serious forms of the disease (4). A role of pre-existing and cross-reacting antibodies, from endemic coronaviruses, that recognize proteins from SARS-CoV-2 is discussed and a phenomenon termed antibody-dependent enhancement (ADE), which is linked to existing antibodies, might be one of the reasons for life-threatening symptoms occurring during later stages of COVID-19 (5,6).

An assay capable of detecting antibodies against endemic coronaviridae, such as OC43, 229E, and NL63, will help to understand a possible role of existing antibodies against these human coronaviruses during COVID-19. Available systems are using recombinant antigens to detect viral protein-directed antibodies in serum or plasma samples. This approach is not only economical but also makes the generation of large reagent batches feasible, allowing for the generation of vast numbers of assays required for systematic patient screening (7,8). Here, we employ a novel way of building a serologic assay system to detect and characterise anti-SARS-CoV-2 antibodies. The approach utilised is based on the classical Western blot procedure, which has been modified to be run as a throughput assay system. The use of the complete pathogen proteome for antibody detection has been employed since the late 1970s (9) and subsequently proven to be useful for the identification of proteins recognized during the humoral immune response. In lysates, prepared from infectious virus particles, not only all possible viral proteins are present and can be probed in one assay, but the use of authentic antigens should enable the detection of antibodies recognising relevant protein modifications present in the naturally occurring pathogen.

The DigiWest procedure, which is employed here, is a variant of the classical Western blot. It addresses the most obvious disadvantages of Western blotting, namely its low throughput, high antigen consumption and poor reproducibility. In the DigiWest, the assay signal is generated on microspheres rather than on a membrane, thus allowing the use of fast and standardised assay protocols on the Luminex platform. Due to the inbuilt possibility of multiplexing, multiple antigens from different viruses can be probed at the same time, enabling the set-up of semi-quantitative seroreactivity screens.

## Materials and Methods

### Patients and blood samples

A total of 263 pre-existing and de-identified serum samples was used for assay development. Ethical approval was granted from the Ethics Committee of University Hospital Tübingen; samples from 193 SARS-CoV-2 polymerase chain reaction (PCR) positive individuals (179/2020/BO2) and of 18 self-reported negative samples were collected (179/2020/BO2). A self-reported healthy serum sample (n=1) and self-reported convalescent serum after SARS-CoV-2 infection (n=2) were obtained at the NMI under the guidelines of the local ethics committees (495/2018/BO2). Sample collection for each donor was performed approximately three to eight weeks after the end of symptoms and / or negative virus smear. In addition, samples from healthy donors obtained from Central BioHub before 8/2019 were used as negative controls (n=49).

### SARS-CoV-2 virus lysate

To prepare SARS-CoV-2 virus lysate, the supernatant of infected human Caco-2 cells was purified. Briefly, Caco-2 cells were infected 1:10 - 1:500 with clinical isolate 200325_Tü1. 48 hours post infection the supernatant was collected, centrifuged and frozen. 900 µL of supernatant was added to 200 µL 20 % sucrose and centrifuged for 90 min at 4 °C and 14000 rpm. The supernatant was discarded and a PBS washing step was done, followed by another centrifugation step. The supernatant was discarded and the viral pellet was re-suspended in 25 µL of LDS sample buffer (Life Technologies) and heated for 5 min at 95 °C.

### Multiplex serum reactivity test *via* DigiWest

Whole viral protein lysates from 229E, OC43, and NL63 (ZeptoMetrix Corp) and from SARS-CoV-2 were used for DigiWest as described. Briefly, viral protein lysates were used for gel electrophoresis and Western blotting using the NuPAGE system. Membranes were washed with PBST (0.1 % Tween-20, PBS) and membrane-bound proteins were biotinylated by adding 50 µM NHS-PEG12-Biotin (Thermo Fisher Scientific) in PBST for 1 h. After washing in PBST, membranes were dried overnight. Subsequently, the Western-Blot lanes were cut into 96 strips of 0.5 mm width and were transferred to a 96-well plate (Greiner Bio-One). For protein elution, 10 µL of elution buffer was added to each well (8 M urea, 1 % Triton-X100 in 100 mM Tris-HCl pH 9.5) The protein eluates were diluted with 90 µL dilution buffer (5 % BSA in PBST, 0.02% sodium azide). Neutravidin-coated MagPlex beads (Luminex) of a distinct colour ID were added to the protein eluates and binding was allowed overnight; 500 µM PEG12-biotin in PBST was added to block remaining Neutravidin binding sites. The bead containing fractions were pooled and thereby the original Western blot lanes were reconstituted. Beads were washed in PBST and resuspended in store buffer (1 % BSA, 0.05 % azide, PBS). The generated bead-set represents the proteomes of the four coronaviruses (SARS-CoV-2, OC43, 229E, NL63) and reactivity against all proteins can be tested in one assay.

For serum incubation, 5 µL of the bead mix were equilibrated in 50 µL serum assay buffer (Blocking Reagent for ELISA (Roche) supplemented with 0.2 % milk powder, 0.05 % Tween-20 and 0.02 % sodium azide, 25 % Low Cross buffer (Candor Bioscience), 25 % IgM-reducing agent buffer (ImmunoChemistry). Serum assay buffer was discarded and 30 µL of diluted patient serum (1:200 in serum assay buffer) was added and incubated for 2 hours at RT on a shaker. After washing in PBST, 30 µL of Phycoerythrin labelled anti-human IgG secondary antibody (diluted 1:200 in serum assay buffer; Dianova) was added and incubated for 45 min at 23 °C. The beads were washed twice with PBST and readout was performed on a Luminex FlexMAP 3D.

The DigiWest analysis tool was used to assess serum reactivity against the viral proteins (10). Virus protein-specific peaks were identified and average fluorescence intensity (AFI) values were calculated by integration of peak areas.

To detect the nucleocapsid of SARS-CoV-2 a commercial antibody was used (Sino Biologicals; 40143-R019). Incubation was performed as described previously (11).

### Statistical analysis

Sensitivity and specificity for each assay were calculated using the results of the PCR-testing as the gold standard. Concordance was calculated using Cohen’s Kappa with 95 % confidence intervals (CI) (12). Correlation was calculated using Spearman’s r with 95 % CI. For determining the dynamic range, a sigmoidal, 4-parameter logistic regression was used to fit the data and interpolate the dilution factor at the cut-off signal. All statistical analyses were performed using GraphPad Prism 8 or R studio (version 1.3.959).

## Results

### DigiWest for detecting serum antibodies against SARS-CoV-2

The DigiWest procedure is a variation of the Western blot that brings this classical approach into significantly higher throughput (9). Here, we used the method for size dependent separation of virus proteins reflecting the whole proteome and their subsequent immobilization on microspheres in order to adapt it to serum analysis. As a first step for detecting, serum antibodies recognizing viral proteins, lysates from infectious SARS-CoV-2 virus particles were prepared in SDS-PAGE loading buffer. DigiWest was performed as described using 0.5 µg of virus protein and Luminex microspheres sufficient to run 100 assays were generated. Detection of total protein on the loaded DigiWest beads (**Fig. 1A**) showed characteristic protein bands for the lysate.

**Figure 1:**
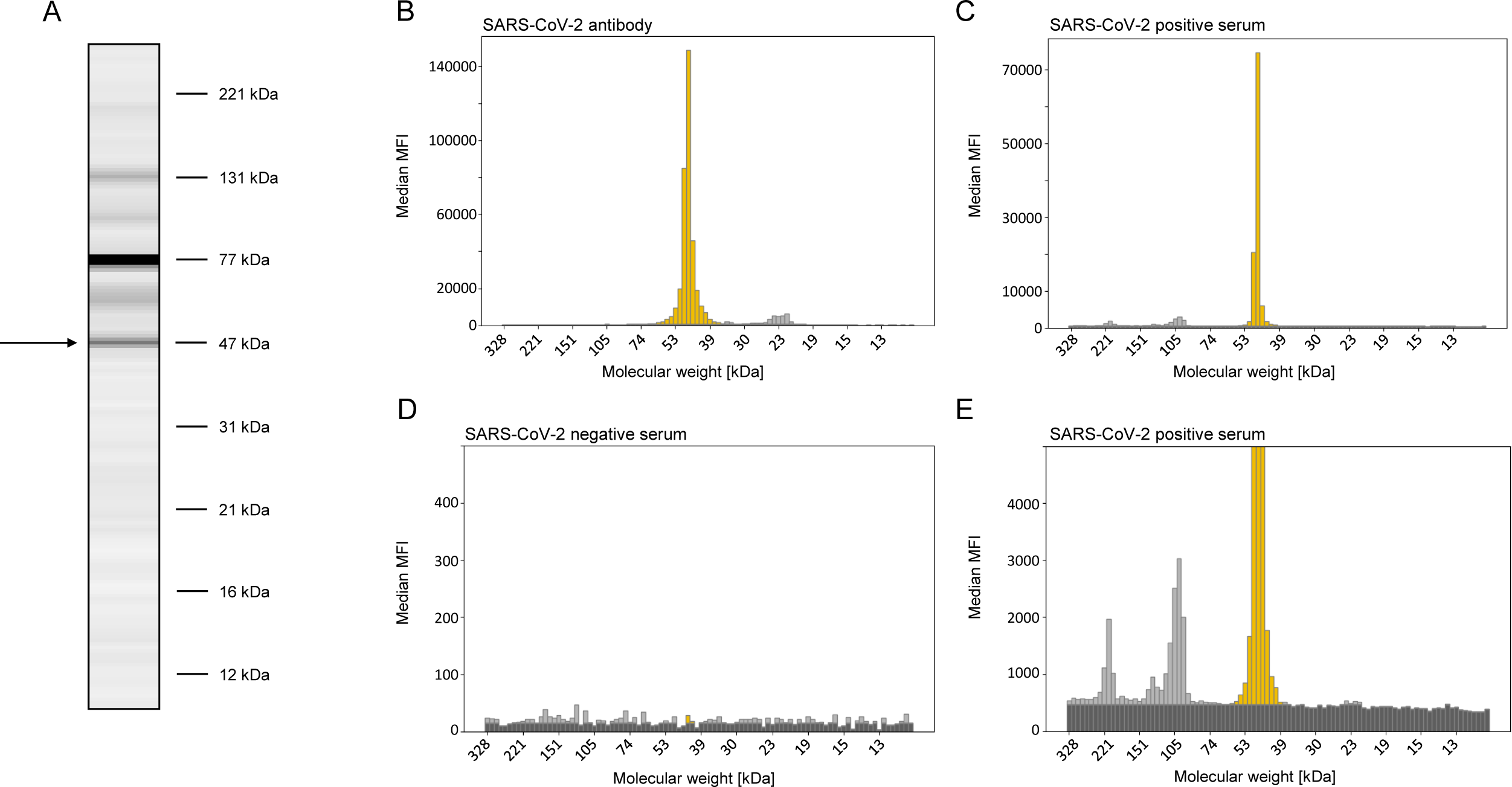
Protein detection on SARS-CoV-2 virus lysate loaded DigiWest beads. Virus proteins were size separated by the Digiwest procedure and transferred to microspheres. In (**A**) a total protein stain of the separated proteins is shown; data are represented as Western-Blot mimic (10) thereby resembling a SDS-Page lane. The marked protein band corresponds to the viral nucleocapsid protein. An anti-SARS-CoV-2 nucleocapsid antibody detects this protein at the expected molecular weight (47.2 kDa) (**B**). Serum from a SARS-CoV-2 PCR positive patient reacts with the same protein (47.2 kDa) giving high fluorescent intensity (**C**), whereas in a negative serum no peaks are detected (**D**). In a fraction of positive sera, additional peaks at 100 kDa and 211 kDa are detected (**E**, detailed view of **C**).

In the next step, human sera were diluted 1:200 in an optimized and modified serum assay buffer (see Materials and Methods) and incubated with the DigiWest microspheres. For SARS-CoV-2 negative samples, no or very low signals were obtained (**Fig. 1D**). High signals were detected from COVID-19 convalescent sera. Most sera showed their main peak of reactivity at 47 kDa, i.e. the size corresponding to the SARS-CoV-2 nucleocapsid protein (**Fig. 1C**). Using an antibody generated against the SARS-CoV-2 nucleocapsid, a prominent peak at 47.2 kDa (**Fig. 1B**) was detected, which is consistent with the expected size of the protein. For a subset of serum samples, additional peaks corresponding to the spike protein are detected (**Fig. 1E**). Assay background was found to be variable, but since the determined signal intensities only consist of the peak area, reliable values were calculated using the DigiWest evaluation tool (10). Since reactivity against the nucleocapsid protein was consistently found in COVID-19 convalescent sera, these values were used for describing SARS-CoV-2 seroreactivity.

### Multiplexed DigiWest for detecting serum antibodies recognizing different human coronaviridae

To expand the assay and to cover human endemic coronaviruses, virus lysates from the two alpha coronaviruses 229E and NL63 and from the beta coronavirus OC43 were processed as described above, and equivalent DigiWest assays were established and combined into one assay system. When using sera from SARS-CoV-2 negative individuals, seroreactivity against viral proteins was found for a large fraction of tested samples. As for the SARS-CoV-2 DigiWest assay, the main serological activity for the different viruses was detected at a molecular weight that corresponds to nucleocapsid proteins. To prove that the detected proteins are indeed the nucleocapsids of the different coronaviruses, we produced recombinant versions of the nucleocapsid proteins of all tested viruses. We used the purified proteins in a different DigiWest experiment and compared the obtained signals with the signals obtained from the whole virus lysate DigiWest (**Fig. 2A**). In the whole virus lysate, the observed molecular weight of SARS-CoV-2 nucleocapsid protein was 47.2 kDa with a calculated molecular weight of 45.6 kDa, for OC43 nucleocapsid protein it was 53.1 kDa (calculated 49.3 kDa), for 229E nucleocapsid protein it was 45.4 kDa (calculated 43.5 kDa) and for NL63 it was 42.1 kDa (calculated 42.3 kDa) and thereby in good agreement with the expected values. The DigiWest using recombinant proteins (**Fig. 2B**) confirmed the molecular weights. A small set of 12 sera was used to detect seroreactivity on virus lysates and on the recombinant nucleocapsid proteins. Correlating signal was detected and this confirmed that the detected reactivity is directed against the nucleocapsid proteins.

**Figure 2:**
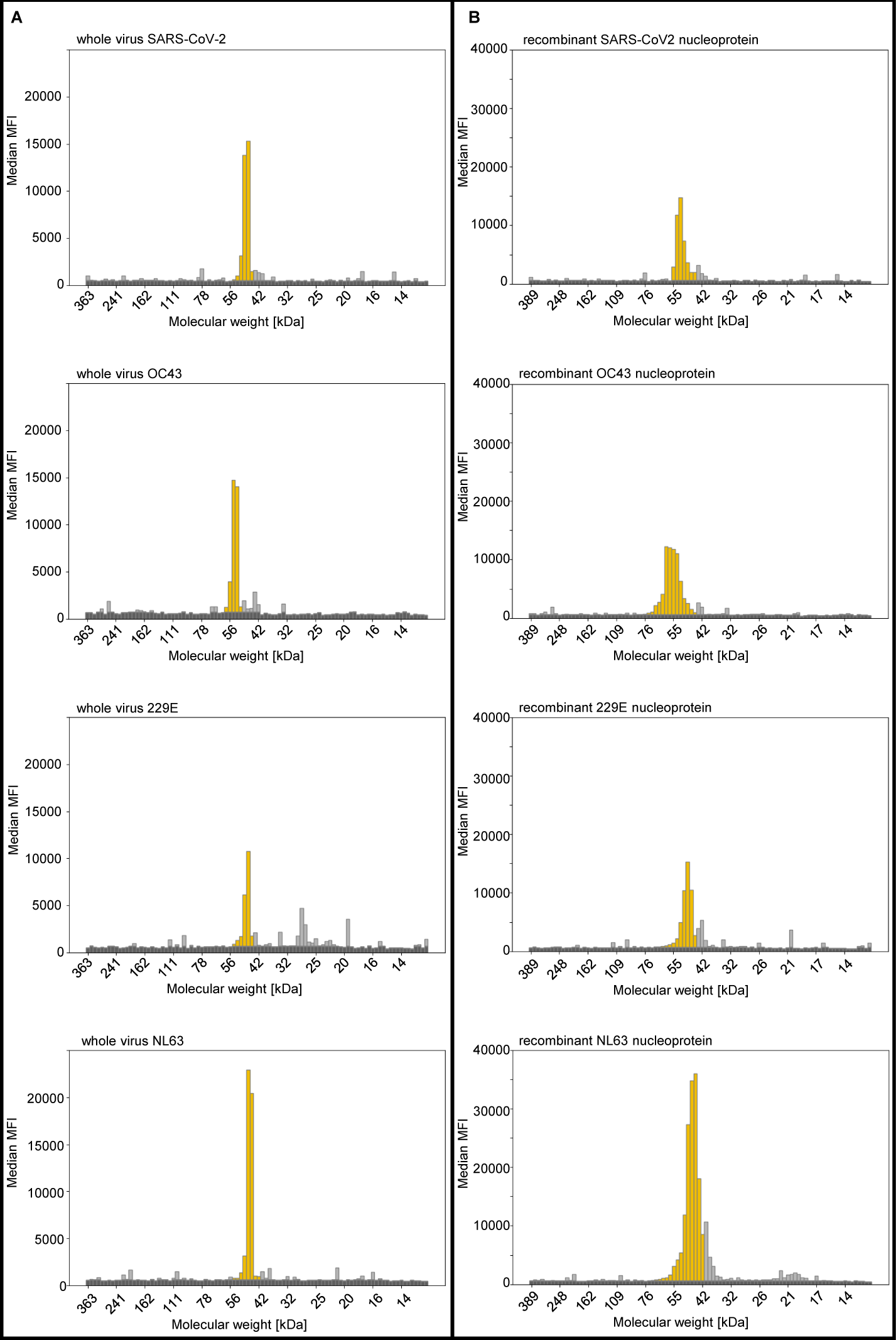
Multiplexed detection nucleocapsid protein from SARS-CoV-2, OC43, 229E, and NL63. Reactivity of a patient serum was tested on whole virus lysates of the different coronavirus types **(A)** and on recombinant nucleocapsid proteins of the different viruses **(B)** using multiplexed DigiWest assays. The used SARS-CoV-2 positive serum shows antibody reactivity on whole virus lysates for (i) SARS-CoV-2, (ii) OC43, (iii) 229E, and (iv) NL63 **(A)**. In **(B)** the same serum is incubated with a DigiWest bead-set loaded with recombinant nucleocapsid from (i) SARS-CoV-2, (ii) OC43, (iii) 229E, and (iv) NL63. As for the whole virus lysates antibody reactivity is observed; for SARS-CoV-2 a peak at 47.2 is kDa detected, for the endemic Coronaviridae OC43, 229E, and NL63 peaks at the molecular weights at the respective sizes of 53.1, 45.4 and 42.1 kDa are found.

### Evaluation of the characteristics of the SARS-CoV-2 serological assay

To characterize the performance of the DigiWest, we used the final multiplexed assay now comprising virus lysates of SARS-CoV-2, 229E, OC43 and NL63 to screen a set of characterized samples (13). Among the analyzed sera, there were 195 SARS-CoV-2 PCR positive specimens, 49 pre-pandemic samples and 19 self-reported negative samples. The complete data set, including a graphical representation of the assay signals in a Western-blot-like format, is available online in Supplemental Data 1. To define the assay cut-off for SARS-CoV-2 seropositivity, 68 non-infected control samples were employed. The highest signal value detected in this group was 1598 AFI. In a second step, the lowest value of all SARS-CoV-2 PCR-positive specimens still above this intensity (1968 AFI) was defined as a seropositive for SARS-CoV-2. The mean of these two measurements was calculated and defined to be the cutoff for seroconversion (1783 AFI). Using this value, an assay specificity of 100 % was found. Furthermore, 26 / 195 (13.3 %) samples from SARS-CoV-2 PCR positive specimens showed no seroconversion, yielding a sensitivity of 87.2 %. These fundamental characteristics of the newly established assay system are comparable to published values for different commercially available SARS-CoV-2 immunoassays (14,15).

To demonstrate the dynamic range of the serologic DigiWest assay, a SARS-CoV-2 positive serum was serially diluted with a negative serum (**Fig. 3**). Good signal linearity was seen in the dilution curve and seropositivity was detected down to a serum dilution of 1:5000.

**Figure 3:**
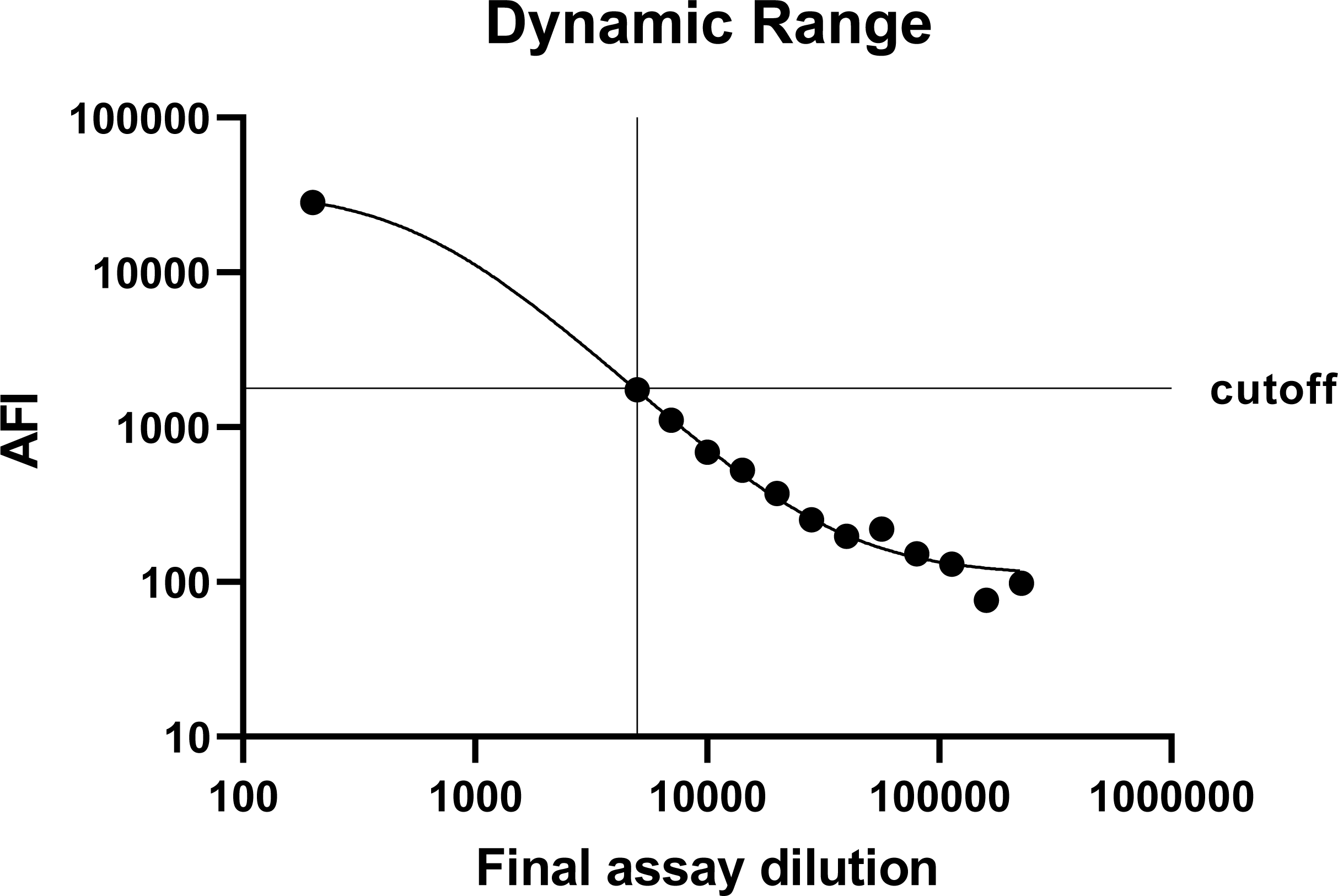
Dynamic range of the DigiWest serological assay. The serum of a SARS-CoV-2 positive patient was diluted in serum of a SARS-CoV-2 negative donor (serial dilution,13 steps ranging from 1:25 to 1:1131). The mixture was further diluted 1:200 in serum assay buffer and the immunoassay was performed. Shown are the final dilutions of positive serum (X-axis) and the resulting average fluorescence intensity (AFI). Logistic regression was performed using a sigmoidal fit and 4-parameter logistics. (Bottom 110.0; Top 34406; IC50 2.962; HillSlope -1.411; logIC50 0.4716).

For closer evaluation of the assay performance, we reanalyzed the complete sample set using the (i) Elecsys® anti-SARS-CoV-2 assay (Roche Diagnostics), (ii) ADVIA Centaur® SARS-CoV-2 (Siemens Healthcare Diagnostics) (16), (iii) EUROIMMUN SARS-CoV-2 IgG ELISA and (iv) EUROIMMUN SARS-CoV-2 IgA ELISA test systems. Further information on the assay procedures are provided in the Supplemental Data 2.

Concordance (Cohen’s kappa) and Correlation (Spearman’s r) analysis were performed and the different assay characteristics were compared and visualized (**Fig.4A-D**). Concordance of DigiWest vs Roche was found to be 0.9429 (95 % CI; 0.90-0.98 **Fig.4A**) and for DigiWest vs Siemens, Cohen’s kappa was 0.8860 (95 % CI; 0.83-0.94 **Fig.4B**). Concordance of DigiWest vs Euroimmun IgG was calculated in two ways: if the borderline results were considered positive, Cohen’s kappa was found to be 0.9102 (95 % CI; 0.86-0.96), if considered negative, the concordance was 0.8799 (95 % CI; 0.82-0.94 **Fig.4C**). When comparing the DigiWest based IgG detection with the Euroimmun based IgA test and borderline results were considered positive, a value of 0.7498 (95 % CI; 0.67-0.83) was found. If the borderline results were considered negative, Cohen’s kappa was found to be 0.7518 (95 % CI; 0.67-0.83).

Correlation analysis utilizing Spearman’s r revealed a positive correlation of all investigated assays (**Fig. 4D**). The highest correlation for DigiWest was found with the Roche system (Spearman’s r= 0.91; 95 % CI; 0.89-0.93). Spearman’s r for DigiWest and Siemens was found to be 0.87 (95 % CI; 0.83-0.90). Spearman’s r for DigiWest and Euroimmun IgG and IgA was calculated at 0.87 (95 % CI; 0.84-0.90) and 0.78 (95 % CI; 0.72-0.82). The highest overall correlation was found between Siemens and Euroimmun IgG (Spearman’s r = 0.94; 95 % CI; 0.92-0.96), the lowest overall correlation was found between Euroimmun IgA and Roche (Spearman’s r = 0.72; 95 % CI; 0.66-0.78). For the calculation of sensitivity and specificity, all assays were compared based on the SARS-CoV-2 PCR status. The determined values for all employed systems are listed in **Table 1**.

**Table 1:**
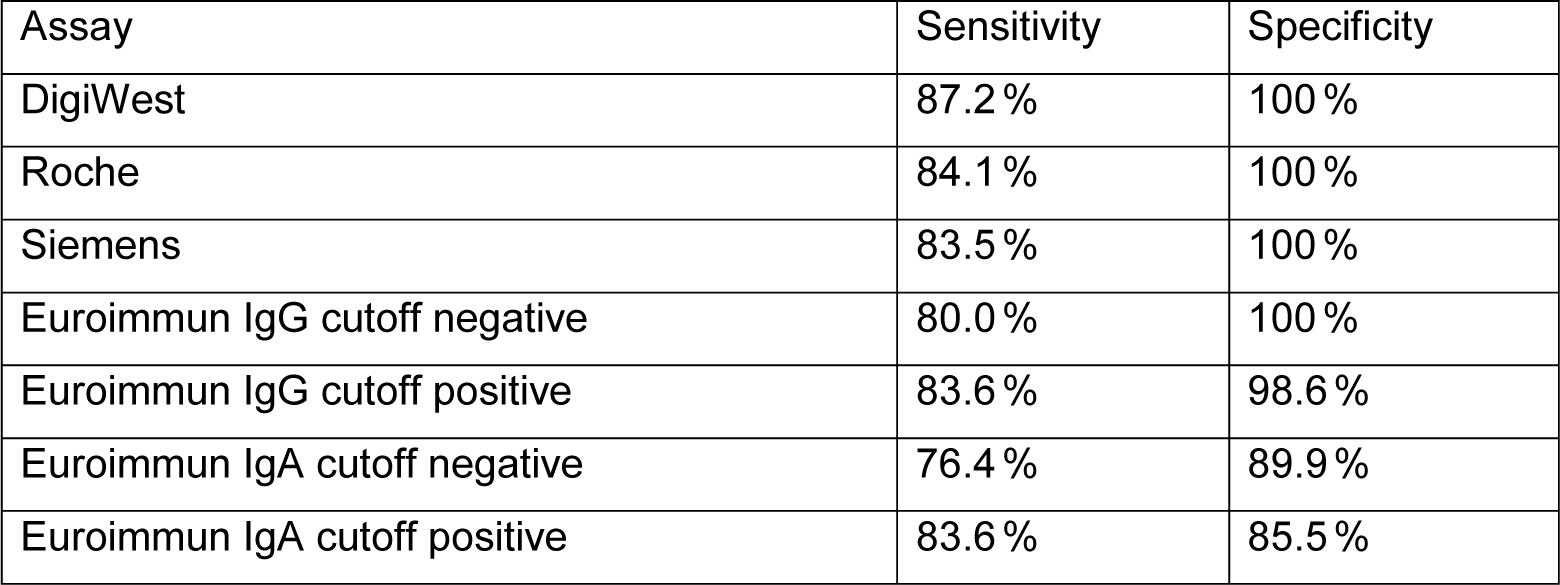
Sensitivity and Specificity for all employed assay systems.

**Figure 4:**
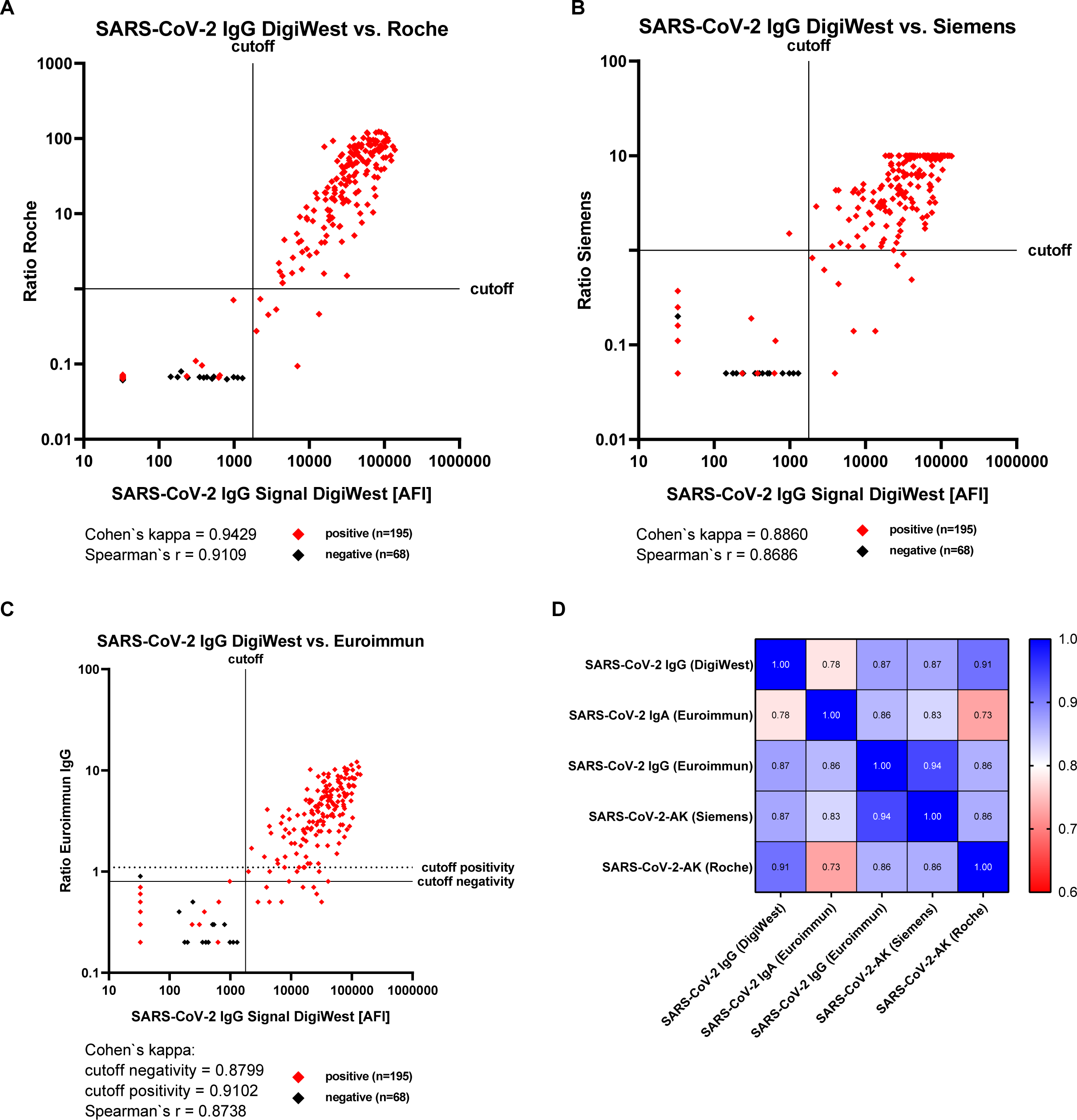
Comparison of the DigiWest seroconversion assay with commercially available SARS-CoV-2 assays. Concordance (Cohen’s kappa) and correlation coefficients (Spearman’s r) of DigiWest data and the commercially assays from Roche (**A**), Siemens (**B**) and Euroimmun IgG (**C**) were calculated and are shown below the plotted data; cutoff values are depicted as a black line in the scatter plot. For the Euroimmun IgG two different kappa values were calculated; when borderline results (as defined by the manufacturer) were considered positive, kappa was 0.9102. If the borderline results were considered negative, the concordance for Euroimmun IgG was 0.8799. In (**D**) the correlation coefficients (Spearman’s r) between all used assays are shown in a heatmap. The highest value (Spearman’s r = 0.91) for DigiWest was found for the Roche system, the lowest value (Spearman’s r = 0.78) for DigiWest vs Euroimmun IgA shown of Spearman’s r values.

### Multiplexed detection of antibodies against SARS-CoV-2, OC43, 229E and NL63

By integrating DigiWest assays for 229E, OC43 and NL63 into the detection system for SARS-CoV-2, concomitant detection of the presence of antibodies binding to antigens derived from the different Coronaviridae becomes possible. In the analyzed sample set, reactive antibodies against all endemic coronaviruses were detected with high frequency (Supplemental Data 1). To estimate the reactivity against the other human endemic coronaviruses, a provisional cutoff for OC43, 229E and NL63 was defined at same value as determined for SARS-CoV-2 (1783 AFI). For Sars-CoV-2 negative sera, 82.4 % showed reactivity against OC43 nucleocapsid, 95.6 % against 229E and 100 % against NL63. For SARS-CoV-2 positive samples, the numbers were 79.5 % against OC43, 99 % against 229E and 98.5 % against NL63. The overall reactivity was 80.2 % against OC43, 98.1 % against 229E and 98.9 % against NL63. Despite the high frequency of antibodies directed against the endemic coronaviruses OC43, 229E, and NL63 in SARS-CoV-2 negative sera, no recognition of SARS-CoV-2 proteins was observed in these samples. This directly translates into the high specificity of the SARS-CoV-2 assay system and reveals that only minor or no cross-reactivity of existing antibodies with the SARS-CoV-2 nucleocapsid protein exists. The correlation analysis between all coronaviruses (including SARS-CoV-2) showed values ranging from 0.03 to 0.75 (**Fig. 5**). The highest correlation was observed for antibodies recognizing the nucleocapsid protein of 229E and NL63 with a Spearman’s r of 0.75 indicating a possible cross-reactivity.

**Figure 5:**
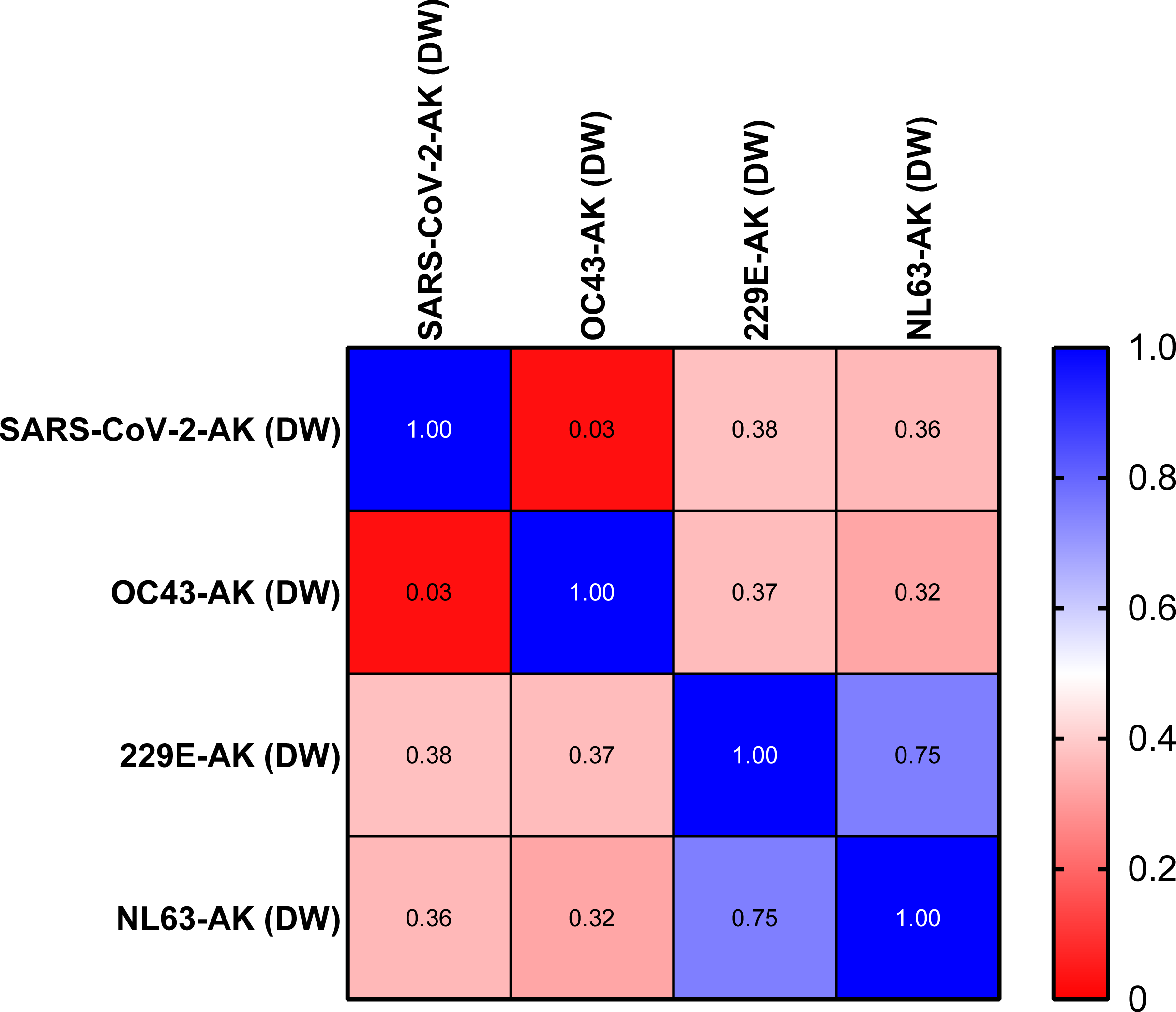
Spearman’s rank correlation of SARS-CoV2 and endemic coronavirus types in the serological DigiWest assay. Data generated for SARS-CoV-2, OC43, 229E and NL63 were used for correlation analysis and Spearman’s rank coefficients were calculated for assay pairing. Results are displayed as heatmap of Spearman’s r values. A high correlation (Spearman’s r 0.75) was found between NL63 and 229E indicating cross-reactivity.

## Discussion

The use of authentic proteins from clinically relevant pathogens as antigens for antibody detection is a classical method for identifying an individual immune response (17). While the approach has distinct drawbacks, *e. g*. the need for isolation of large amounts of pathogen and poor assay reproducibility when using different protein batches, it also provides substantial advantages. Modifications only found in the authentic proteins are present in antigen preparations and therefore the identification of reactive antibodies against these possible pathogen-derived antigens should be feasible. In addition, the generation of protein extract from pathogens of different strains is often technically uncomplicated and fast. This may turn out to be especially useful when a comparative analysis of antigen preparations from closely related pathogenic agents is of interest. Such an analysis may facilitate the identification of relevant cross-reacting antibodies directly on a wide variety of antigenic structures. These advantages may help to set up systems that take an unbiased approach to characterising the humoral immune response and may allow the identification of cross-reacting antibodies.

Here we describe the set-up of such an assay system using protein extracts prepared directly from infectious SARS-CoV-2 virus particles. The employed DigiWest procedure is an immunoblot system that closely resembles the classical Western blot procedure. After SDS-PAGE based protein size separation, proteins are immobilized on polystyrene microspheres and assay read-out is performed on the Luminex assay platform. 10 micrograms of protein are sufficient to generate batches of assay material for thousands of serum analyses; this directly translates into good assay reproducibility. In addition, the use of the Luminex platform for read-out allows for a high assay throughput without the need for producing recombinant proteins. As in Western blotting, the assay gives direct information on the size of the recognized proteins, and often antigenic proteins can be directly identified. When using COVID-19 convalescent sera, a specific antibody response to a protein of 47 kDa corresponding to the nucleocapsid protein of SARS-CoV-2 was recurrently seen. Reactivity against other viral proteins was present in individual serum samples, yet the nucleocapsid protein was identified as the major antigen in this assay. The observed low seroreactivity against the spike protein is due to the fact that reduced and denatured proteins were employed for the DigiWest and that these protein forms are not recognized by most of the anti-spike antibodies (data not shown).

For the detailed evaluation of the performance of the newly developed assay for detecting anti SARS-CoV-2 antibodies a set of more than 250 well-characterized sera was employed, which were mainly taken from a clinical study on T-cell response after SARS-CoV-2 infection (13). By using 4 different serological assays that are in use in clinical routine labs we showed high concordance (Cohen’s Kappa 0.88-0.94) between all systems. The detected specificity as compared to the PCR results was found to be ranging from 85.5 % to 100 %, with the DigiWest, the Roche and the Siemens systems approaching 100 %. Sensitivity was found between 76.4 % and 87.2 %, with the DigiWest reaching the highest score here. This demonstrates high standards for all tested assays. Interestingly, the highest concordance (0.94) was found between the Siemens assay system and the Euroimmun IgG assay, with both assays mainly detecting the spike protein. Nearly the same Kappa value was calculated for the Roche and the DigiWest system, both of which use the nucleocapsid protein as the detected antigen. The Euroimmun IgA showed slightly different assay characteristics, which is most likely due to the fact that it is the only assay that exclusively detects IgA immunoglobulins. Yet, no principle differences in assay characteristics were observed and all assays showed high quality.

Antibodies against endemic coronaviruses are frequently found in human individuals (18). These viruses cause mild diseases and are associated with approximately 20 % of the common colds (19,20). However, when comparing the sequences of the virus genome, the degree of similarity between the SARS-CoV-2 and these viruses is astonishingly high (21). This similarity has led to speculations that antibodies against these endemic viruses may also possess protective properties against SARV-CoV-2 (22). The presence of these antibodies might explain the vastly diverse courses of disease. Therefore, the DigiWest assay system was expanded by using lysates from alpha coronaviruses 229E and NL63 as well as the beta coronaviruses OC43, thus enabling the detection of serum antibodies recognizing antigens from these four coronaviruses from one specimen in one assay. The implementation of these assays directly followed the method used for SARS-CoV-2 and seroreactivity against the nucleocapsid protein was frequently found for these coronaviruses.

As expected a very high rate of infection for all of the coronaviruses was found, yet no indication of cross-reactivity to the SARS-CoV-2 proteins was seen in pre-pandemic and other SARS-CoV-2 negative samples. This is in contrast to the described T-cell response that can be triggered by peptides derived from the SARS-CoV-2 nucleocapsid (13,23).

Since the described assay was optimized for specificity, the employed assay conditions were highly stringent to avoid the occurrence of false positive signals. Modifications of the assay that use less stringent conditions assays could be used to detect cross-reactivity of antibodies formed against proteins from endemic coronaviruses to SARS-CoV-2 viral proteins. This could give valuable information on the nature of these frequently found anti-corona antibodies; this work is ongoing.

As a serologic assay system the use of the DigiWest approach is not only novel, but it allows the set-up of a highly specific assay within a very short time frame and it is capable of detecting a wide variety of serum antibodies since all pathogen-derived proteins can be probed in one reaction.

## Data Availability

Supplementary material is available online.

## Author Contributions

All authors confirmed they have contributed to the intellectual content of this paper and have met the following 4 requirements: (a) significant contributions to the conception and design, acquisition of data, or analysis and interpretation of data; (b) drafting or revising the article for intellectual content; (c) final approval of the published article; and (d) agreement to be accountable for all aspects of the article thus ensuring that questions related to the accuracy or integrity of any part of the article are appropriately investigated and resolved.

## Acknowledgement

This work has received funding from the European Union’s Horizon 2020 research and innovation program under grant agreement No 101003480 - CORESMA.

## Supplemental Material

Supplemental Material is available online.

